# Optimizing multi-domain hematologic biomarkers and clinical features for the differential diagnosis of unipolar depression and bipolar depression

**DOI:** 10.1101/2022.04.26.22274241

**Authors:** Jinkun Zeng, Yaoyun Zhang, Xiang YuTao, Sugai Liang, Chuang Xue, Junhang Zhang, Ya Ran, Songfang Huang, Fei Huang, Luo Si, Tao Li, Wei Deng

## Abstract

There is a lack of objective features for the differential diagnosis of unipolar and bipolar depression, especially those that can be easily accessible in practical settings. Some studies have shown that unipolar and bipolar depression have different associations with hematologic biomarkers and clinical features such as the age of onset. However, none of them have used these features for differential diagnosis. We investigated whether biomarkers of complete blood count, blood biochemical markers and clinical features could accurately classify unipolar and bipolar depression using machine learning methods.

1,160 eligible patients were included in this retrospective study (918 with unipolar depression and 242 with bipolar depression). 27 biomarkers of complete blood count,17 blood biochemical markers and 2 clinical features were investigated for the classification. Patient data was split into training (85%) and test set (15%). Using ten-fold cross validation for training, logistic regression (LR), support vector machine (SVM), random forest (RF) and Extreme Gradient Boosting (XGBoost) were compared with feature selection.

We calculated the AUC, sensitivity, specificity and accuracy. The optimal performance was achieved by XGBoost using a combination of selected biomarkers of complete blood count (WBC, PLR, MONO, LYMPH, NEUT Ratio, MCHC, BASO Ratio, LYMPH Ratio), blood biochemical markers (albumin, potassium, chlorine, HCT, calcium, LDL, HDL) and clinical features (disease duration, age of onset). The optimal performances achieved on the open test set were AUC 0.889, sensitivity 0.831, specificity 0.839 and accuracy 0.863. Hematologic biomarkers and onset features seem to be reliable information that could be easily accessible in clinical settings to improve diagnostic accuracy. In addition, we further analyzed the importance of specific blood biomarkers in samples of disease durations <= 3 years and > 3 years. WBC and MONO remained informative across different disease durations. Meanwhile, NEUT, BASO Ratio, HCT and LYMPH, and albumin were more indicative in the short course (<= 3 years), whereas NLR and chlorine were more indicative in the longer course (> 3 years). This may suggest that, given the overall stability of the model, longitudinal changes in biomarkers should be investigated across different disease courses and age groups.

## Introduction

Mood disorder, also known as affective disorder, is a group of significant and lasting mood or emotional changes caused by a variety of reasons. It is mainly manifested by recurring emotional ups and downs, often accompanied by corresponding behavioral and cognitive changes.[1] In the international classification system of mental disorders International Classification of Diseases 10th edition (ICD-10) and the American classification system American Diagnostic and Statistical Manual of Mental Disorders 4th edition (DSM-IV), mood disorders mainly include: major depressive disorder, bipolar disorder, persistent mood disorder and cyclothymic disorder, etc.[2] However, in the new classification standard of the fifth edition of DSM (DSM-V) [3], the mood disorder is regarded as two independent categories, including bipolar and bipolar related disorders, as well as depressive disorder. Accordingly, the ICD-11[4] will also be revised to be synchronized with DSM-V.

Major depressive depression, also called unipolar disorder (UPD), is a serious mental disorder. Its main clinical features include low mood, lack of interest, and loss of pleasure, which are accompanied by loss of appetite, sleep disorders, low self-evaluation, pessimistic and world-weariness. There are also changes in patients’ cognition and behavior.[1,5] In contrast, bipolar depression (BD-DEP) refers to the depressive phase of bipolar disorder.[6] There are many similar clinical manifestations in patients of unipolar and bipolar depression, with depressive mood as the most important manifestation for both of them. Most patients diagnosed with bipolar disorder spend more time in the depressive phase than in the manic phase, and the onset of depression is relatively more common than manic. Moreover, early in the course of the disease, patients with bipolar disorder may not have a clear history of hypomania/manic episodes.[7] There is also a lack of epidemiological features or subliminal symptoms of hypomania/mania that can assist in differential diagnosis.[8-11] Therefore, it is difficult to catch such differential symptoms, which are frequently ignored by both doctors and patients. Other clinical symptom features that are used to distinguish between the two diseases generally yield low performances (in terms of accuracy, sensitivity, specificity and other indicators) [12-14], which are not sufficient to be used in clinical practice. This makes it difficult to distinguish between unipolar and bipolar depression during the phase of disease onset, especially bipolar disorder II. Surveys have shown that only 20% of patients with bipolar depression can be correctly diagnosed at the first episode, and it takes an average of 10-15 years for patients to get a correct diagnosis.[15-17]. BD-DEP is often misdiagnosed as UPD, which resulting in mistreatment with an unopposed antidepressant. Antidepressants are often ineffective for treating BD-DEP and may cause detrimental consequences such as treatment-emergent hypomania/mania, rapid cycling, or increased suicidality. On the other hand, despite the clinical manifestations of UPD and BD-DEP are very similar, the clinical outcomes and treatment options of the two are completely different. For UPD, antidepressant drugs are used clinically to treat depressive symptoms and prevent their recurrence. For BD-DEP, in addition to treating depressive symptoms, it is also necessary to prevent (hypo)manic episodes, so it is mainly treated with mood stabilizers and/or atypical antipsychotics.[18, 19]

Considering that most patients with bipolar disorder have a depressive phase for the first time [20], and depressive episodes remain to be predominant throughout the course of the disease (the number of depressive episodes is about 3 times the number of manic episodes) [21, 22], it is necessary to study the pathological basis of UPD and BPD, to find objective biomarkers to distinguish between them and establish a high-performance differential diagnosis model. Related works proposed to use different domains of biomarkers, such as serum levels [23], MRI [24-26] and cognitive functioning [10], in statistical and machine learning methods for differential diagnosis of UPD and BPD. Particularly, there are extensive studies and meta-analysis regarding longitudinal associations between inflammatory biomarkers such as CRP/IL-6 and UPD, and between blood based protein biomarkers and BPD. However, the problem with current approaches is that it is hard to access such biomarkers in real-life practice. On the other hand, biomarkers of common blood count and blood biochemical markers can be conveniently accessible in clinics, through low-cost and reproducible tests that can be easily calculated from a blood sample collected under simple laboratory conditions.

Previous works have attempted to find the association between biomarkers of common blood count and mood disorders. For example, the white blood cell (WBC) count, a non-specific inflammatory marker, is usually measured as part of the complete blood count (CBC) panel. The association between WBC subtypes and affective disorders has already been established in previous studies.[27-30] In addition, neutrophil-to-lymphocyte ratio (NLR), platelet-to-lymphocyte ratio (PLR), and monocyteto-lymphocyte ratio (MLR) have recently been proposed as inflammatory markers. These biomarkers appear associated with mood disorders, supporting the inflammatory hypothesis underlying the etiopathogenesis of these conditions.[27, 31] Up to the present date, several studies have examined the usefulness of NLR, PLR, and MLR as potential biomarkers to differentiate BPD from UPD.[31,32]

As for the biochemical markers, the human body’s antioxidant defense system includes two categories of antioxidants, enzymatic and non-enzymatic. Although it is difficult to measure the levels of enzymatic antioxidants such as superoxide dismutase and catalase, the detection of non-enzymatic antioxidants such as albumin are included in daily liver and kidney function tests and can be conducted conveniently. Albumin and other non-enzymatic antioxidants can be used to monitor the antioxidant level of the body.[33] Previous works showed that the concentrations of plasma albumin and other non-enzymatic antioxidants in patients with depression are low; the analysis of major depression, manic and control groups also showed that the albumin in the major depression group is lower than that in the mania group, and both were lower than those in the control group.[cite] Furthermore, association between serum uric acid and depressive symptoms reported in PHQ-9 were identified,[34] changes in blood lipid levels were also reported to be associated with schizophrenia, UPD and BPD.[35] However, to the best of our knowledge, no study has compared BPD with UPD in terms of blood biochemical indicators.

Despite unipolar and bipolar depression have different associations with clinical features such as the age of onset and hematologic biomarkers, none of previous works have used such information for differential diagnosis. It is urgent to establish objective features that can be easily accessible in practical settings and develop accurate differential diagnosis models for UPD and BPD.[36, 37] In this study, we took the initiative to build and validate an automated diagnostic algorithm, based on hematologic biomarkers and clinical features, to classify UPD and BPD in practical settings.

## Materials and methods

### Participants

This was a naturalistic, retrospective, cross-sectional study. All the patients were from Hangzhou Seventh People’s Hospital and completed blood related examinations. Data from 2018.01-2021.06 was collected. All data available in that period was analyzed. Only the baseline complete blood count and biochemistry tests of the first entry for each patient from inpatient care units was used for analysis. Usually, the first blood tests are done next day after admission to our units when the patients were after 12 h of fasting. Thus, we have assumed that the most patients that we included in this study were in acute phase of their disorder. We mainly included patients between the ages of 18 and 65 years who were diagnosed with BPD and UPD based on the ICD-10 criteria. Patients were grouped under diagnostic criteria as unipolar depression (F32 and F33 according to ICD-10) and bipolar depression (F31.3-F31.5 according to ICD-10). As a general rule, most patients with coexisting severe somatic diseases (e.g., acute autoimmune and inflammatory diseases, renal failure, cancer or other), which may significantly affect various blood parameters, are not included in our study. Thus, we assume that observed results are mostly related to psychiatric conditions.

The initial number of patients with UPD is 1 318, the initial number of patients with BPD is 507. As illustrated in Figure 1, 107 patients of UPD with psychotic symptoms, and 113 patients of BPD with psychotic symptoms were removed from the cohort; 290 patients of UPD and 155 patients of BPD with missing values of any biomarkers were removed from the cohort. Finally, 921 and 239 patients left as participants in the study, respectively.

**Figure 1.**
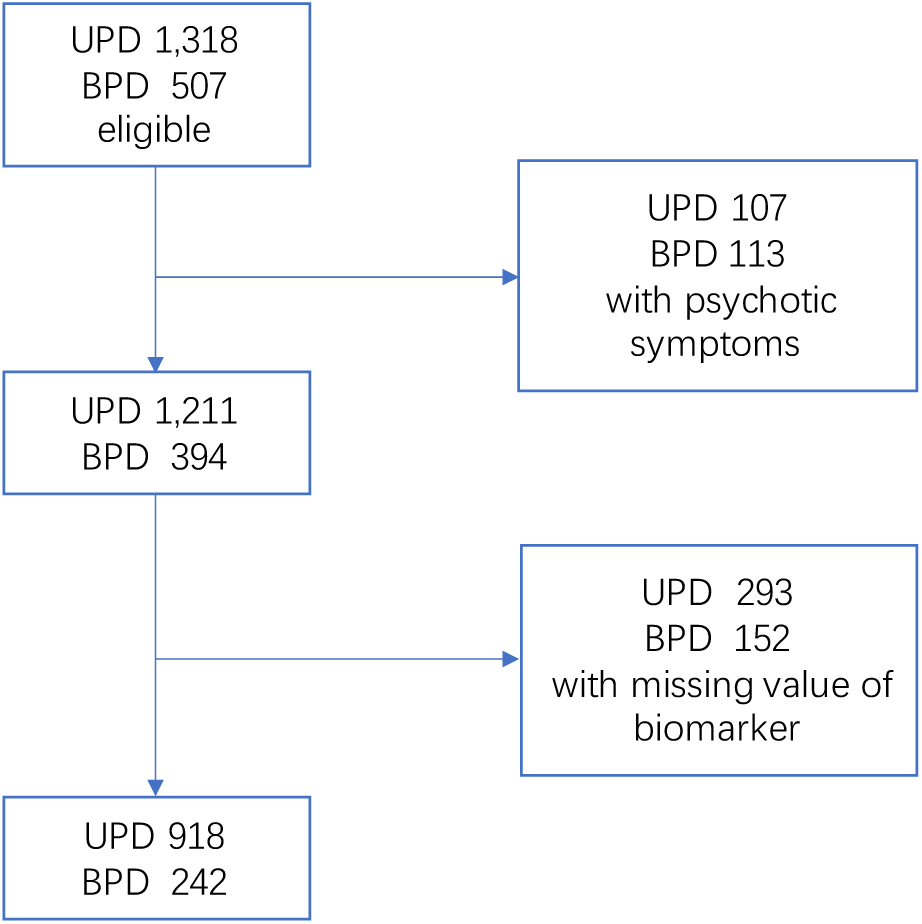
Flow of participants in the study. Patients with psychotic symptoms and missing values of biomarkers were excluded from the participants. As a result, a total of 921 UPD patients and 239 BPD patients participated in the study.

### Biological and clinical data acquisition

#### Clinical data acquisition

Basic socio-demographic and clinical features were obtained through electronic medical record System including age, gender, diagnosis, age of onset and duration of the disease.

#### Blood data acquisition

A complete blood count was performed to evaluate red blood cell (RBC) count, hemoglobin (HGB), hematocrit (HCT), mean corpuscular volume (MCV), mean corpuscular hemoglobin (MCH), mean corpuscular hemoglobin concentration (MCHC), red blood cell distribution width-coefficient of variation (RDW-CV), neutrophils, lymphocytes, monocytes, eosinophils and basophils, and platelets. biochemistry tests were performed to evaluate calcium, chloride, potassium, magnesium, sodium, phosphorus, globulin ratio, albumin, total protein, globulin, creatinine, urea nitrogen, low-density lipoprotein (LDL), high density lipoprotein (HDL), triglycerides, and total cholesterol. Blood samples were taken in the morning (between 7 and 9 a.m.) of the first day of hospitalization, after 12 h of fasting, from a forearm vein. For each patient, about 3 ml of blood was collected in hemogram tubes containing EDTA. After collecting blood samples, complete blood count was determined using Sysmex XN-3000 Automated Hematology Analyzer (Sysmex, USA). Blood biochemistry was analyzed using Siemens automatic biochemical analyzer-XTP.

### Statistical analysis

Statistical analyses were conducted by the IBM SPSS Statistics 22 (IBM SPSS, Turkey). The comparison of the sex distribution between the two groups was performed usingχ2 test. Comparisons including age, age of onset and total disease duration between the two groups were performed using a two-tailed two-sample t test. Unless specified otherwise, the significance of all tests was set to p < 0.05.

### Machine learning process

Figure 2 illustrates our study design of classification-based differential diagnosis between unipolar and bipolar depression: given clinical data sources of both unstructured text in chief complaint and structured tables, features of three domains are extracted from all subjects, including clinical features, common blood biomarkers and blood biochemical markers. Then, two feature selection algorithms, analysis of variance (ANOVA) [38] and SHapley Additive exPlanations (SHAP) [39], were adopted to select most informative features. After that, four classification algorithms were used for differential diagnosis including support vector machine (SVM), logistic regression (LR), random forest (RF), and Extreme Gradient Boosting (XGBoost) methods. Models were trained using 10-fold cross-validation and further evaluated on the open-test dataset. The final output is a diagnosis of unipolar or bipolar depression.

**Figure 2.**
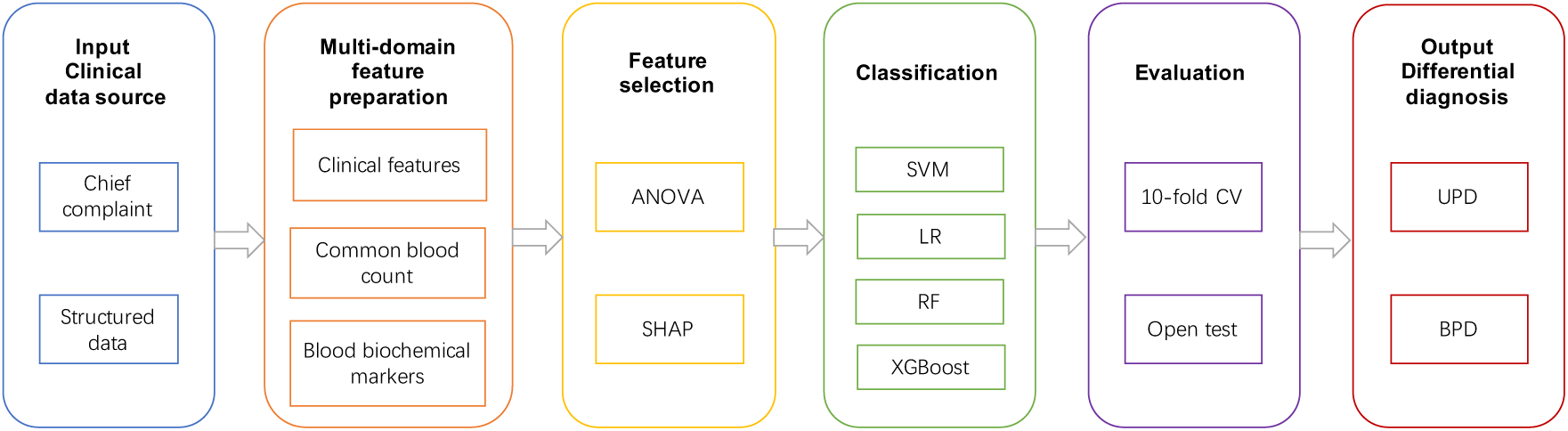
Study design of classification-based differential diagnosis between unipolar and bipolar depression. From clinical data sources of both unstructured text in chief complaints and structured tables, features of three domains were extracted, including clinical features, common blood biomarkers and blood biochemical markers. Then, two feature selection algorithms, ANOVA and SHAP, were adopted to select the most informative features. After that, four classification algorithms were used for differential diagnosis including support vector machine (SVM), logistic regression (LR), random forest (RF), and Extreme Gradient Boosting (XGBoost) methods. Models were trained using 10-fold cross-validation and further evaluated on the open-test dataset. The final output was a diagnosis of unipolar or bipolar depression.

#### Feature preparation

After a thorough survey of potential features for differential diagnosis between UPD and BPD identified in previous research and a discussion with physicians about the important information available in the hospital EHR system, features of three domains were prepared (Figure 3) and examined in this study:

**Figure 3.**
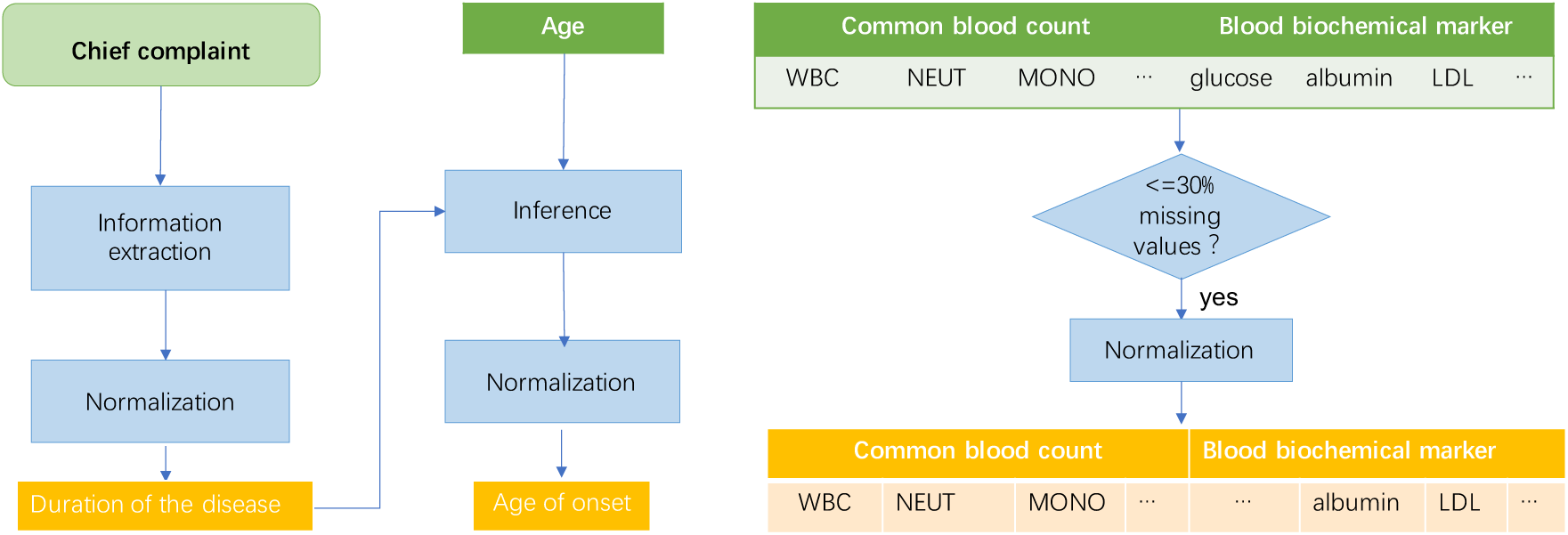
Workflow of feature preparation:two clinical features, duration of the disease and age of the disease onset, were extracted and calculated based on chief complaints; common blood biomarkers and blood biochemical markers with more than 30% missing values were removed, remaining 37 biomarkers normalized into z-scores.

(1) two clinical features including duration of the disease and age of the disease onset. The chief complaints in clinical notes usually started with the major symptoms of patients and the total durations. Therefore, duration of the disease was first extracted from the free text of chief complaints using regular expressions of temporal patterns. For example, from the text of “*Sleep disturbances for 3 years 2 months*.”, “*3 years 2 months*” was extracted and considered as the duration of the disease. Next, the age of the disease onset could be inferenced by subtracting the duration from the age of the patient. The disease duration was normalized in the unit of year, and the age of onset was normalized into four groups – 1 for age <= 20, 2 for age <=40 and >20, 3 for age <=60 and >40, and 4 for age<=65 and >60.

(2) 27 common blood biomarkers including a) biomarkers of the leukocyte system (WBC markers): WBC, MONO, MONO ratio, NEUT, NEUT ratio, BASO, BASO ratio, EO, EO ratio, LYMPH, LYMPH ratio; b) biomarkers of the erythrocyte system (RBC markers): RBC, HCT, MCV, RDW-CV, HGB, MCH, MCHC, RDW-SD; c) biomarkers of the platelet system (platelet markers): PLT, PDW, MPV, P-LCR; d) Blood count-related inflammatory markers: NLR, PLR, MLR, which can be used to evaluate the inflammatory state of the body.

(2) 17 blood biochemical markers including a) electrolyte markers: calcium, chloride, potassium, magnesium, sodium, phosphorus; b) protein markers: globulin ratio, albumin, total protein, globulin; c) markers of kidney function including creatinine, urea nitrogen; d) marker of blood sugar - glucose; e) markers of blood lipids: LDL, HDL, triglycerides, and total cholesterol.

Abbreviations of blood biomarkers used in this study and their full names are listed in Supplement Table 1. Biomarkers with more than 30% missing values were removed (magnesium, globulin ratio, total protein, nitrogen, glucose, triglycerides, total cholesterol). The remaining 37 biomarkers were normalized into z-scores and went through feature selection in this study.

#### Feature selection

In order to select effective features and improve the disease classification performance, two feature selection algorithms, analysis of variance (ANOVA)[38] and SHapley Additive exPlanations (SHAP)[40], were used on each classifier. ANOVA measures the relevance of features to the categories (i.e., UPD and BPD) by determining whether their means come from the same distribution or not, whereas a SHAP value for a feature of a specific prediction represents how much the model prediction changes when we observe that feature.

#### Machine learning methods

Four algorithms were used for classification:

1. LR: LR estimates the parameters of a logistic model; it is a form of binomial regression.
2. SVM: SVM is based on the statistical learning theory of the VC dimension and the structural risk minimization principle. SVMs use kernel functions (such as radial basis kernel functions and linear kernel functions) to project high-dimensional samples into lower dimensions to improve the prediction or classification ability of the model.
3. RM: RM is an ensemble learning method that operates by constructing multiple decision trees during training.
4. XGBoost: XGBoost is a scalable tree-based gradient boosting algorithm. It generates accurate predictions by integrating weak classifiers.

### Evaluation

The dataset was divided into a training set (85%) and a test set (15%) for differential diagnosis evaluation. Different machine learning models were built using the training set, and the parameters were continuously adjusted using ten-fold cross-validation to optimize the models. The final performance of each machine learning algorithm was evaluated using the intended test dataset. Taking BPD as the positive class, standard metrics, i.e., the area under the receiver operating characteristic curve (AUC), sensitivity, specificity and accuracy, were reported to evaluate the classification performance of different models. The AUC usually provides a view of the performance stability. In a general situation, an AUC of 0.90–1.0 is regarded as very high (excellent), of 0.80–0.89 high (good), of 0.70–0.79 moderate (fair), of 0.60–0.69 low (poor), and of 0.50–0.59 as very low (fail or useless).[29] Meanwhile, sensitivity, specificity and accuracy can provide a more objective model assessment from other aspects. Balanced sensitivity and specificity scores were reported based on the ROC (receiver operating characteristic) curves.

In our experiments, machine learning algorithms were implemented using the scikit-learn version 0.24.2 packages. Values of biomarkers were normalized to z-scores using the StandardScaler in scikit-learn. The optimal parameters were selected using GridSearchCV. Performances of using clinical features, blood biomarker features and their combinations were reported, respectively. Moreover, to understand the effectiveness of the differential diagnosis model on patients at early/later stages of the disease course, performances on the samples with disease durations <=3 years and >3 years were also reported, respectively. The dataset was randomly split into a training set and a test set for 1,000 times, and average performances were reported in this study.

### Ethical aspects

The study was carried out in accordance with ethical principles for medical research involving humans (WMA, Declaration of Helsinki). All data were collected anonymously. The study protocol was approved by the Ethical Committee of the Hangzhou Seventh People’s Hospital.

## Results

### Participants

1,160 inpatients of unipolar and bipolar depression were enrolled in the present study, of which 918 were experiencing UPD and 242 were experiencing BPD. The mean (±SD) age of the total sample was 36.86 (±15.04). 771 (66.47%) were females, and 89.31% (N =1136) were employed. As for the total duration of the disease, the mean (±SD) length was 5.18 (±7.80). Other socio-demographic and clinical characteristics are displayed in Table 1.

**Table 1.**
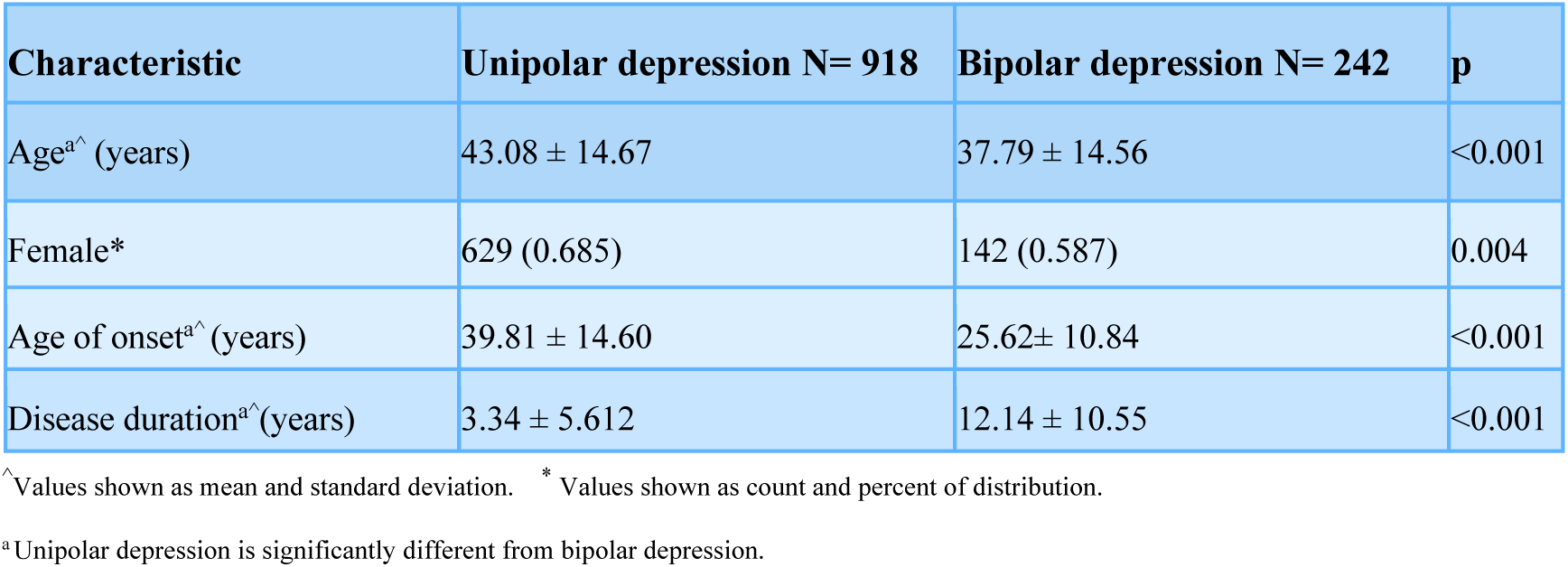
Socio-demographic and clinical characteristics of samples in this study

Figure 4(a) illustrates the cohort size distributions of different durations of UPD and BPD. It is interesting to observe that a majority (∼64%) of UPD patients had a duration <= 3 years. The percentages of patients with UPD (∼20%) and BPD (∼17%) were most close in the duration of 3 - 5 years, from where the percentage of BPD patients increased consistently up to ∼41% in the duration of >10 years.

**Figure 4.**
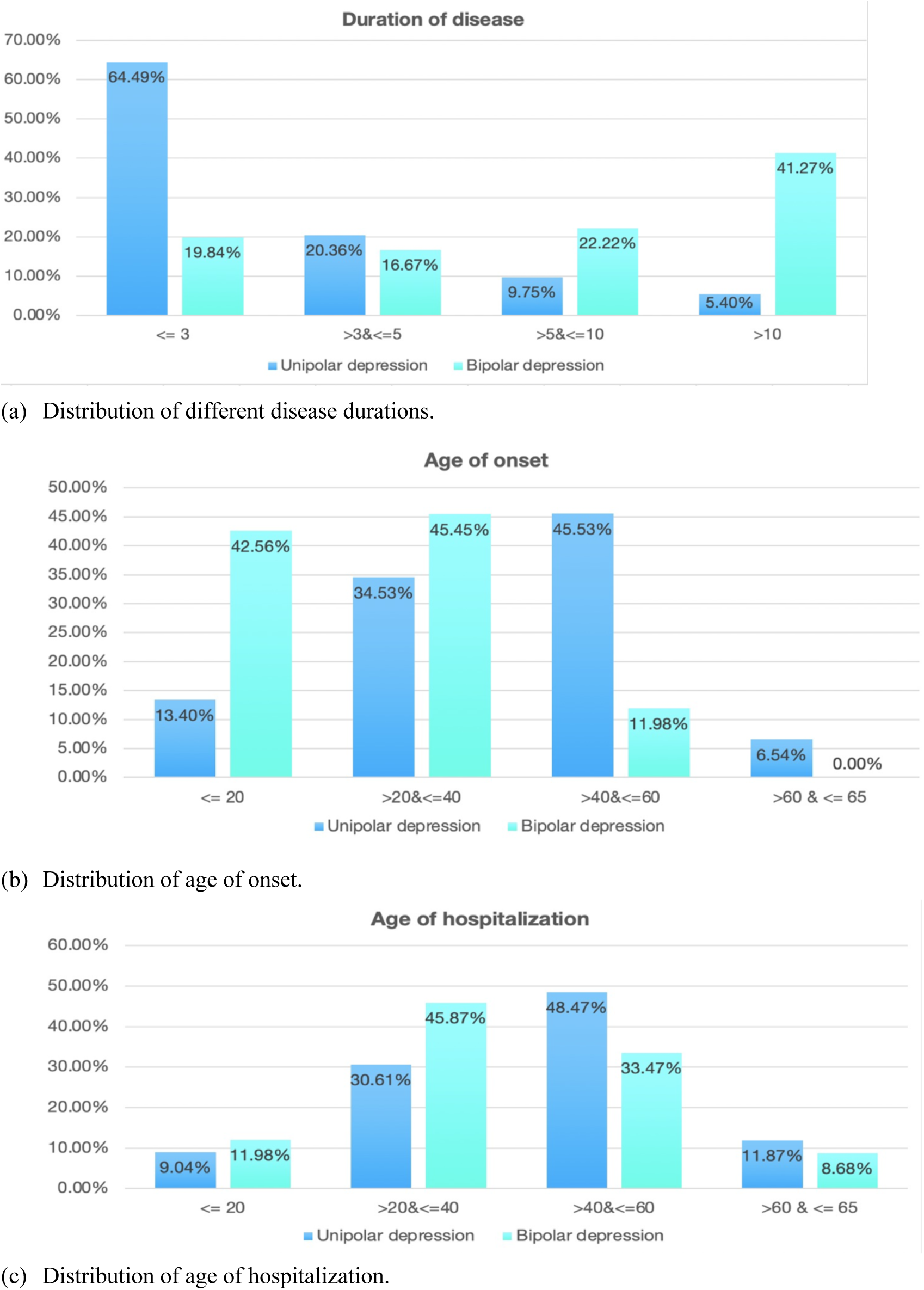
Sample distributions of unipolar and bipolar depression (Unit: year): (a) Distribution of different disease durations. (b) Distribution of age of onset. (c) Distribution of age of hospitalization.

Figure 4(b) illustrates the distributions of ages of onset for UPD and BPD. ∼43% of BPD patients had an early onset <= 20 years old and ∼45% of them had an onset between 20 and 40. In contrast, the majority of onset ages of UPD were between 20 to 60. Taking a look at both Figure 4 and Figure 5, we can find that BPD had a relatively earlier age of onset and a longer duration of disease, while UPD had a relatively later onset of the disease and a shorter duration. Such differences are typical for UPD and BPD, indicating that the cohort used in this study is representative.

**Figure 5.**
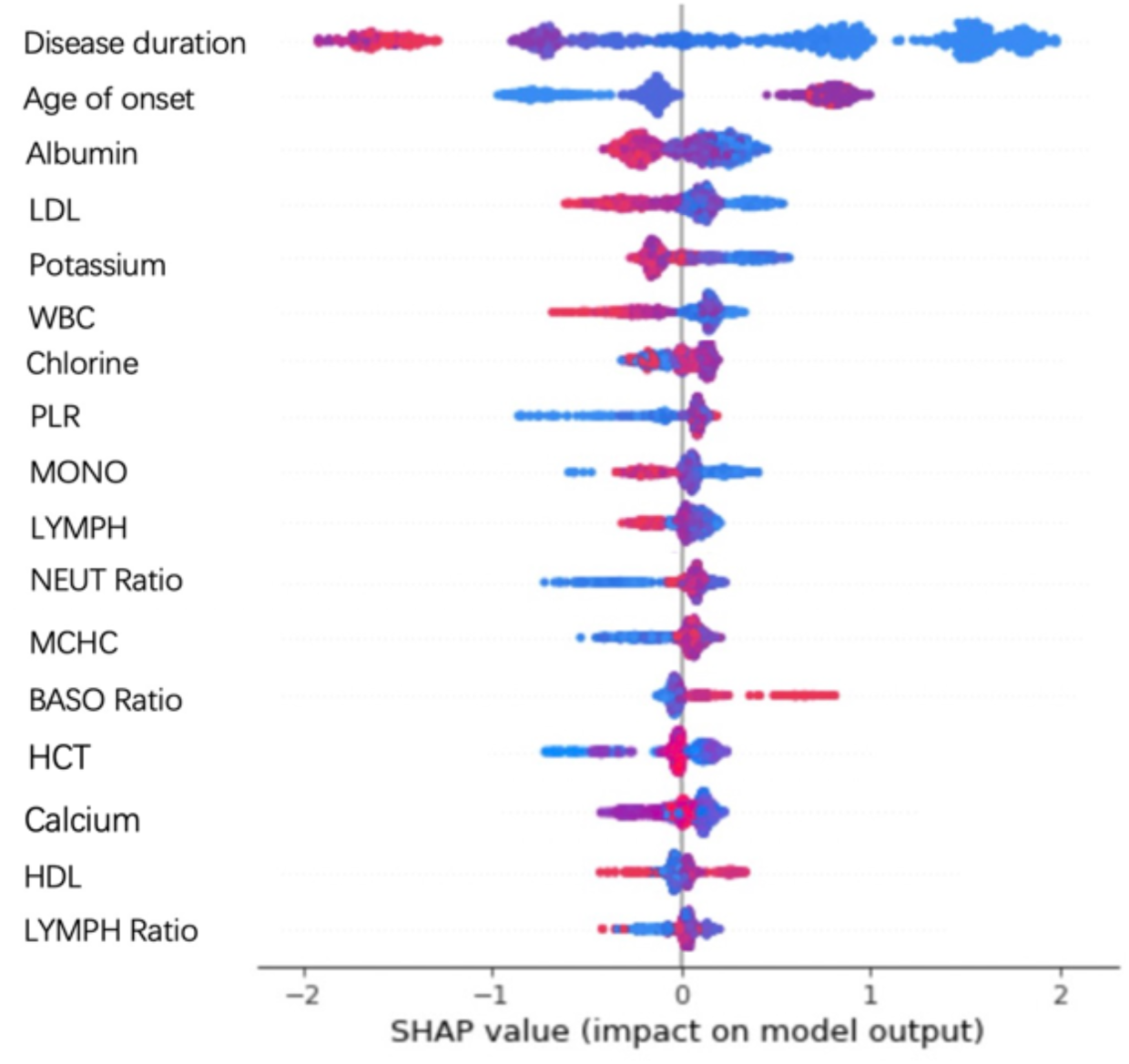
Dot plot of feature importance calculated using the mean SHAP values from running 10-fold cross validation on the training set for 1 000 times. The x-axis in Figure 5 was the SHAP value in units of log odds of BPD. Each row shows the importance of different values of one feature, with red color indicating high feature values and blue color indicating low feature values. The rows were ranked by the overall feature importance vertically. Therefore, disease duration has the strongest drive to the model’s prediction, while LYMPH ratio has a relatively weaker drive. Notably, when points don’t fit together on the line, they pile up vertically to show density.

Following Figure 4(a) and 4(b), Figure 4(c) illustrates the distributions of ages of patient hospitalization for UPD and BPD. The majority patients were between 20 to 60 years old for both diseases (∼79%). Notably, an approximate shape of symmetry could be observed for the age distributions of UPD and BPD patients. The hospitalized patients were relatively younger for BPD (∼12% <= 20, ∼46% between 20 and 40), while the hospitalized patients were relatively older for UPD (∼48% between 40 and 60, ∼12% between 60 and 65).

### Selected features for differential diagnosis

Table 2 lists seventeen features selected by SHAP values for classification in this study, which yielded the optimal performances with XGBoost. To further look into the importance of each feature, a summary plot was drawn with all the SHAP values for a single feature as depicted in Figure 5. The same set of features were selected by ANOVA as the top seventeen features, with different orders of importance. Both clinical features, disease duration and age of onset were considered as effective features. Selected features from the domain of common blood count included four WBC biomarkers - WBC, MONO, NEUT Ratio, and BASO Ratio, two RBC biomarkers – HCT and MCHC, two biomarkers of platelets - LYMPH and LYMPH Ratio, and one biomarker of inflammation - PLR. Six biochemical markers were selected, including three electrolyte markers - potassium, chlorine, and calcium, one protein marker - albumin, and two markers of blood lipids - HDL and LDL. A ranking of the entire feature set based on SHAP values can be found in Supplement Figure 1.

**Table 2.** Clinical features and blood biomarkers selected for classification in this study.

| Domain | Selected features |
| --- | --- |
| Clinical feature | Disease duration, age of onset |
| Common blood count | WBC markers: WBC, MONO, NEUT Ratio, BASO Ratio;<br>RBC markers: HCT, MCHC;<br>Platelet markers: LYMPH, LYMPH Ratio;<br>Biomarker of inflammation: PLR |
| Blood biochemical marker | Electrolyte markers: potassium, chlorine, calcium;<br>Protein markers: albumin;<br>Markers of blood lipids: HDL, LDL |

The x-axis in Figure 5 was the SHAP value in units of log odds of BPD. Each row shows the importance of different values of one feature, with red color indicating high feature values and blue color indicating low feature values. The rows were ranked by the overall feature importance vertically. Therefore, disease duration has the strongest drive to the model’s prediction, while LYMPH ratio has a relatively weaker drive. Notably, when points don’t fit together on the line, they pile up vertically to show density.

### Performance of differential diagnosis

XGBoost using SHAP for feature selection achieved the optimal performances, which were reported in Table 3 and Figure 6(a). Moreover, to understand the effectiveness of the differential diagnosis model on patients at early/later stages of the disease course, performances on the samples with disease durations <=3 years (Table 4 and Figure 6(b)) and >3 years (Table 5 and Figure 6(c)) were also reported, respectively. Performances of other algorithms were reported in Supplement Table 2.

**Table 3.**
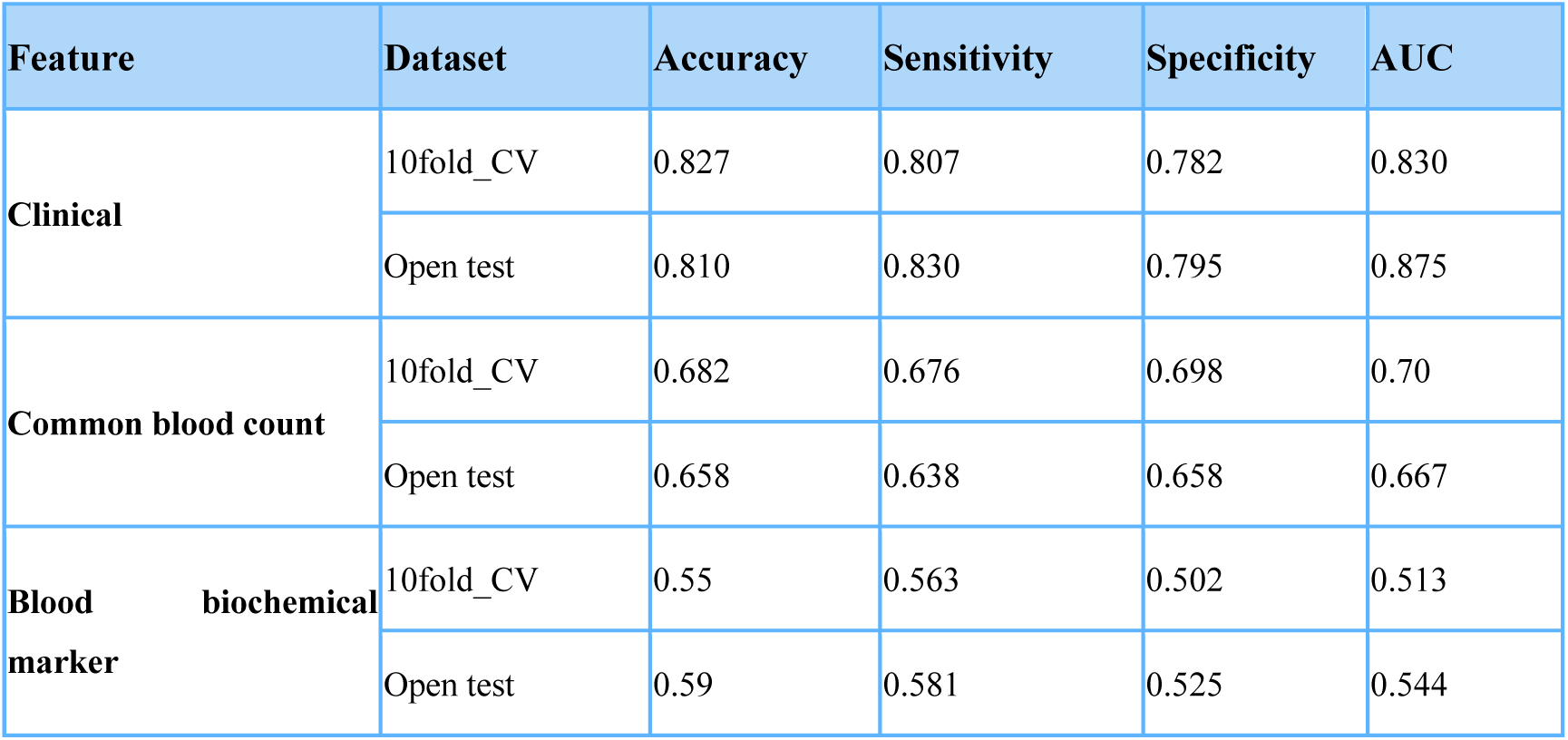
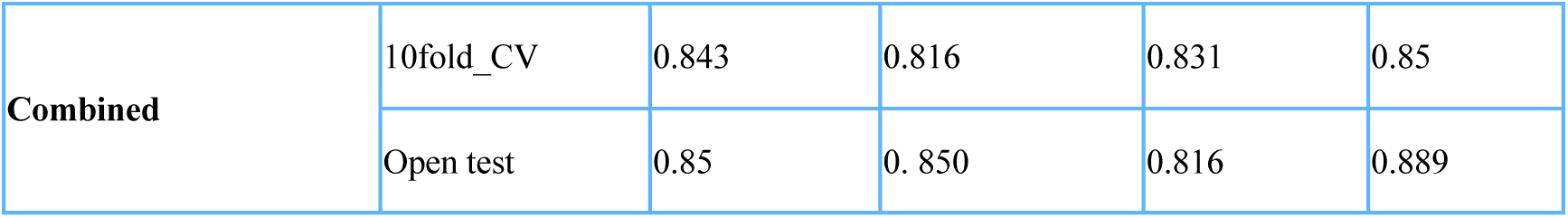
Classification performances of XGBoost using clinical features, hematologic biomarkers and their combination, based on the entire cohort.

**Table 4.**
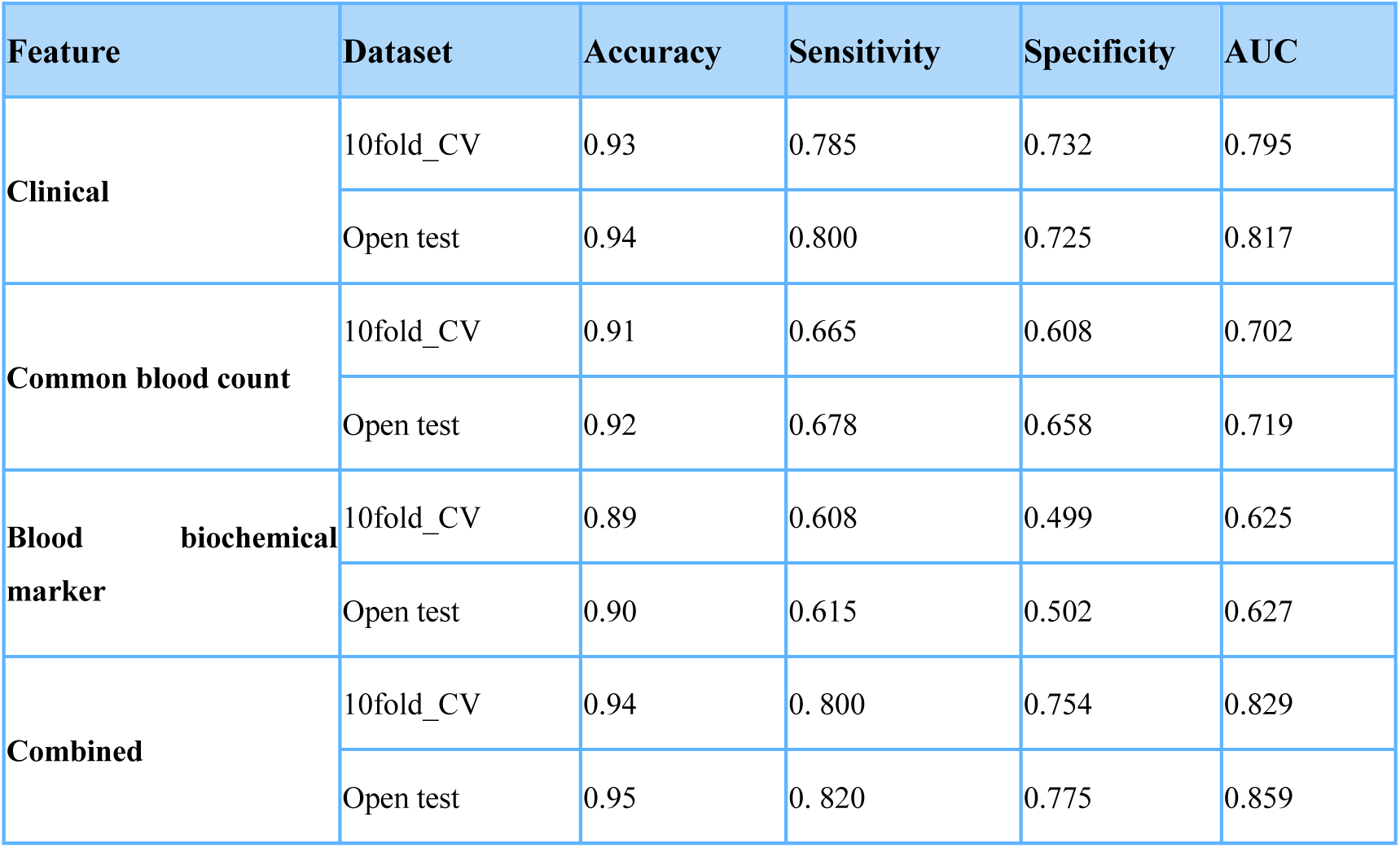
Classification performances of XGBoost using clinical features, hematologic biomarkers and their combination, based on samples of disease duration <=3 years.

**Table 5.** Classification performances of XGBoost using clinical features, hematologic biomarkers and their combination, for samples of disease duration >3 years.

| Feature | Dataset | Accuracy | Sensitivity | Specificity | AUC |
| --- | --- | --- | --- | --- | --- |
| **Clinical** | 10fold_CV | 0.66 | 0.700 | 0.687 | 0.751 |
|  | Open test | 0.71 | 0.712 | 0.692 | 0.768 |
| **Common blood count** | 10fold_CV | 0.66 | 0.700 | 0.517 | 0.669 |
|  | Open test | 0.61 | 0.675 | 0.498 | 0.623 |
| **Blood biochemical marker** | 10fold_CV | 0.47 | 0.632 | 0.292 | 0.505 |
|  | Open test | 0.55 | 0.656 | 0.293 | 0.535 |
| **Combined** | 10fold_CV | 0.70 | 0.789 | 0.664 | 0.783 |
|  | Open test | 0.71 | 0.792 | 0.663 | 0.786 |

**Figure 6.**
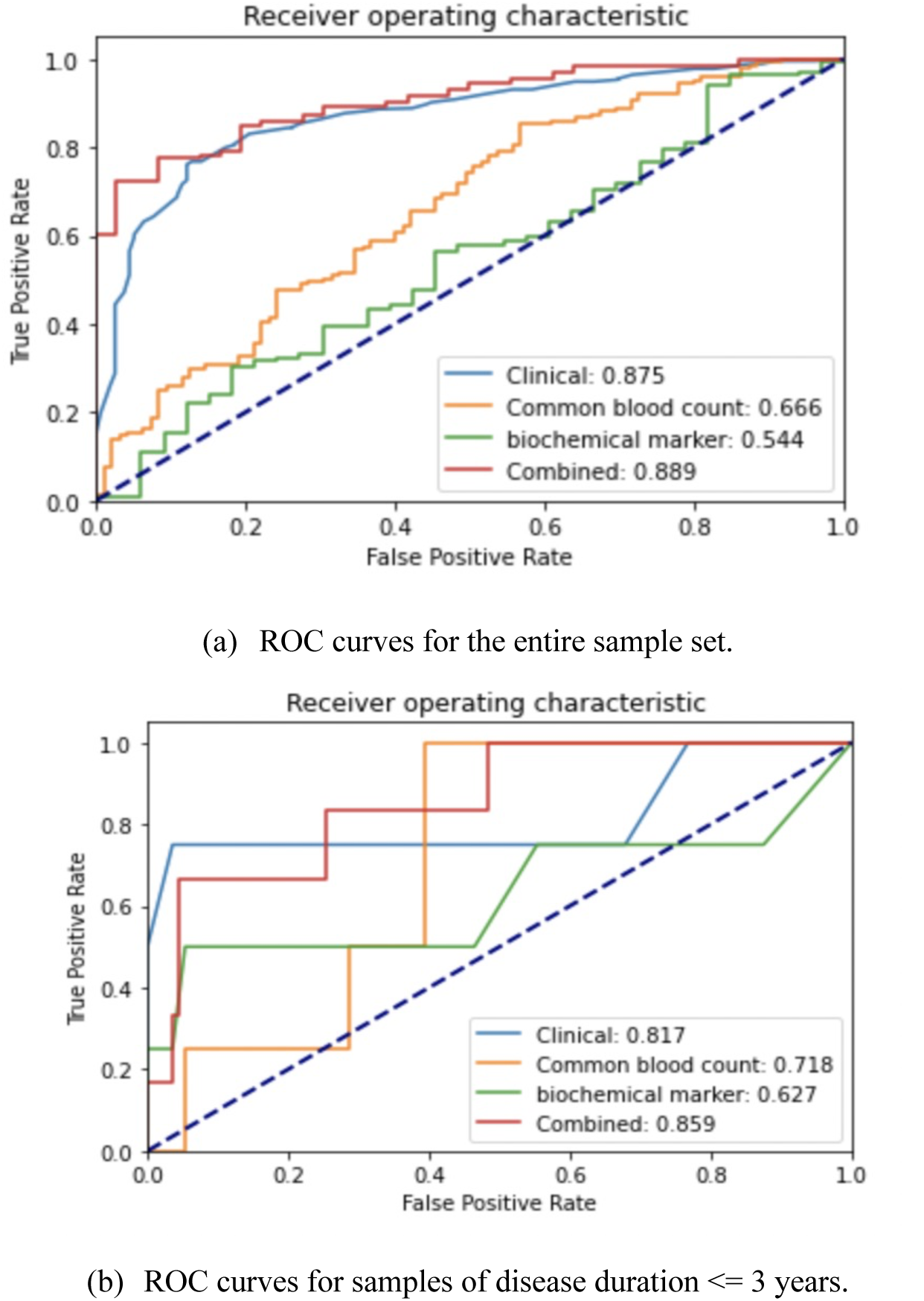
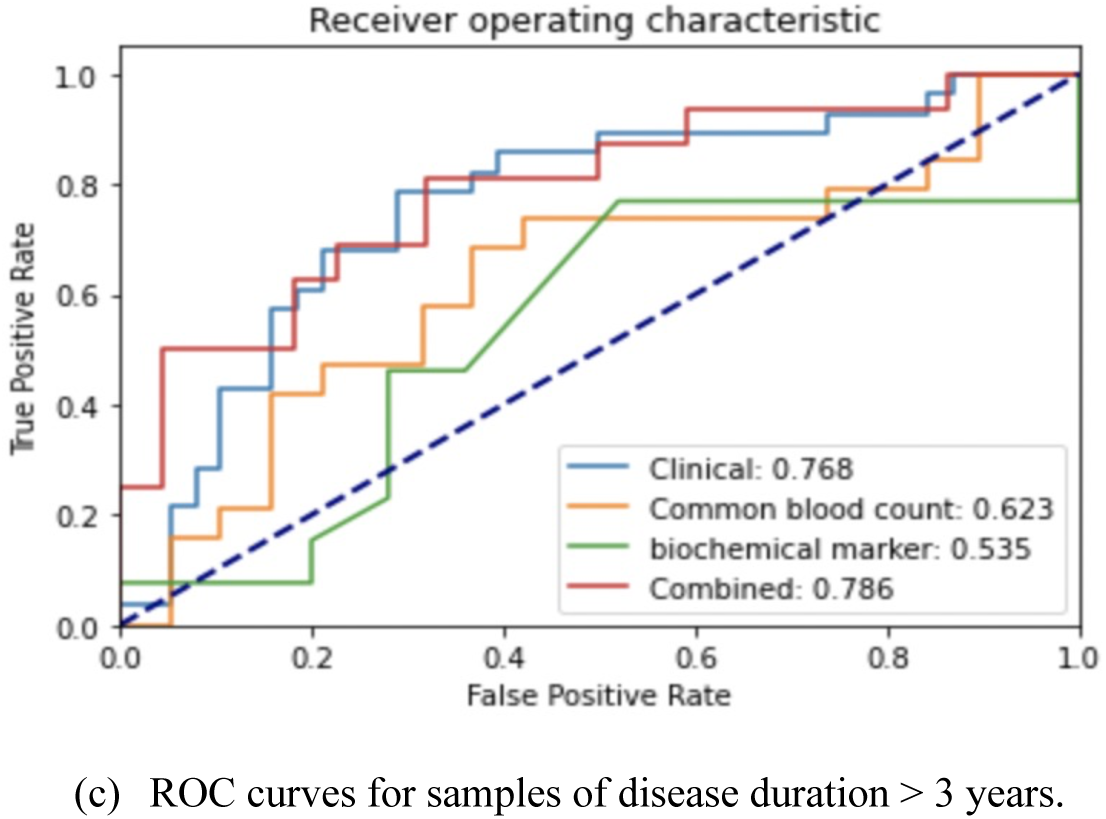
ROC curves of using clinical features, hematologic biomarker and combined features on the open test data, respectively: (a) ROC curves for the entire sample set. (b) ROC curves for samples of disease duration <= 3 years. (c) ROC curves for samples of disease duration > 3 years.

Classification performances of using clinical features, hematologic biomarkers and their combination are displayed in Table 3 and Figure 6(a). Performances on both the training and the test sets are reported. Clinical features produced good AUC (train: 0.830, test: 0.875) and sensitivity (train: 0.807, test: 0830), while biomarkers of common blood count only obtained modest AUC (train: 0.70, test: 0.667). Besides, blood biochemical markers yielded the lowest performances (AUC train:0.513, test: 0.544). Combining all three domains of features got the optimal AUC (train: 0.85, test: 0.889) and sensitivity (train: 0.816, test:0.850). In particular, combined features achieved a boost in the specificity performance (train: 0.831, test: 0.816), which is vital in clinical practice settings. The levels of both AUCs of the clinical features and the combined features could be considered as good.

Classification performances of using clinical features, hematologic biomarkers and their combination for samples of disease duration <=3 years were displayed in Table 4 and Figure 6(b). The overall performances followed the same pattern of performances on the entire samples as shown in Table 4. Clinical features outperformed among the three types of features. Interestingly, the combined features achieved a higher improvement of AUC over the clinical features (Training: 0.795 vs. 0.829; Test: 0.817 vs. 0.859), in comparison with AUC improvement on the entire samples (Training: 0.830 vs. 0.85; Test: 0.875 vs. 0.889). Besides, accuracies on this sub-dataset were relatively higher and did not subject much to the performance changes of the other metric criteria, potentially due to the heavily imbalanced labels (UPD: 681, BPD:50). Similarly, both AUCs of the clinical features and the combined features can be considered as in the level of good.

Classification performances of using clinical features, hematologic biomarkers and their combination for samples of disease duration >3 years were displayed in Table 5 and Figure 6(c). Performances on this subset dropped sharply probably because of the reduced sample size. The optimal AUC was 0.786 produced by the combined features. The AUCs of the clinical features and the combined features could be considered as fair, which may still be helpful to diagnosis in a practical setting.

## Discussion

The clinical manifestations of UPD and BPD are similar, especially during the depressive episodes of BPD.[41] The differential diagnosis of them is based on a comprehensive consideration of medical history, course characteristics, clinical symptoms, and physical, mental and laboratory examinations. In terms of symptom characteristics, the atypical depressive symptoms of patients with BPD are more prominent.[42] Although one or more of the above characteristics can help distinguish UPD from BPD, the current identification performance is not sufficient for practical use.[43] Therefore, it is necessary to study their pathological basis and find objective features to establish a high-performance differential diagnosis model.

As the most common clinical testing methods, common blood count and blood biochemical testing are more accessible and cheaper than genetic and omics-related testing.[44,45] Besides, electroencephalogram and imaging examinations in studies are usually collected in experimental settings and the patient’s coordination to these examinations is relatively lower in practical settings.[46-48] Therefore, previous studies using features from other domains were conducted on relatively small sample sets.[10] [23-26] To the best of our knowledge, this discriminative study of UPD and BPD is the first to combine blood-biological data and data of illness courses, with the largest sample set of 1 160 participants. We developed an integrated framework of machine learning to discriminate patients with UPD from BPD. The main findings of this study are described below: (1) using a combination of biological features and clinical features as input features for the classification, the best performance was achieved, with an AUC of 0.889, a sensitivity of 0.831, a specificity of 0.839 and an accuracy of 0.863. (2) the most discriminative features include selected biomarkers of complete blood count (WBC, PLR, MONO, LYMPH, NEUT Ratio, MCHC, BASO Ratio, LYMPH Ratio), blood biochemical markers (albumin, calcium, potassium, chlorine, HCT, LDL, HDL) and clinical features (disease duration, age of onset).

Studies have found that some epidemiological and symptomatic characteristics help distinguish UPD from BPD.[22] In terms of medical characteristics, the female/male ratio of BPD is lower than that of UPD, the age of first episode is earlier, and the possibility of having a family history of bipolar disorder is higher.[49] Patients with BPD also have the characteristics of rapid depressive episodes, longer duration of illness, and frequent episodes.[50] In addition, patients with BPD have more hospitalizations caused by mental illness and are less responsive to antidepressant drugs.[15] An analysis of samples in this study demonstrated that BPD had a younger age of onset, longer overall disease duration, and a higher proportion of women, which is consistent with existing findings.

Interestingly, among the blood-related indicators, the top ranked features were mainly from the biochemical biomarkers, including albumin, LDL and potassium, instead of biomarkers of common blood count. As mentioned in the introduction, to a certain extent, patients with mood disorders have decreased antioxidant capacity and oxidative stress damage. This study found that albumin and LDL were significantly different in BPD and UPD, which may serve as potential biochemical indicators for their differential diagnosis.[51] This finding also suggests that a patient’s physical condition may have a strong influence on mental state and warrants further study as a next step.

Another interesting finding is that the effective rate (AUC) of the model could reach 0.875 when the clinical features of disease course were used alone, and after adding blood-related indicators, the effective rate increased to 0.889. Notably, previous studies usually constructed cohorts with equal samples of UPD and BPD, which may not reflect their incidence in practical settings. This study measured and reported the differential performances on the original proportion of patients (UPD: 918, BPD: 242). Moreover, this study also examined the performances of samples with different disease durations with different prevalence rates (i.e., <=3 years and >3 years) for the first time, to further understand the effectiveness of the differential diagnosis model on patients at early/later stages of the disease course. The importance of clinical features and blood biomarkers on differential diagnosis were consistent across different disease durations, as illustrated in Table 6. In addition, we further analyzed the importance of specific blood biomarkers in samples of disease durations <= 3 years and > 3 years. WBC and MONO remained informative across different disease durations. Meanwhile, NEUT, BASO Ratio, HCT and LYMPH, and albumin were more indicative in the short course (<= 3 years), whereas NLR and chlorine were more indicative in the longer course (> 3 years). This may suggest that, given the overall stability of the model, changes in biomarkers should be investigated across different courses of disease and age groups.

Limitations and future work:(1) One limitation is that some blood biochemical markers were removed from the original feature set due to more than 30% missing values, which may have the potential to be important features and further improve the performance. (2) Some confounders such as smoking/alcohol status and psychotropic medications may influence the values of blood biomarkers. However, the distribution of such confounders was independent from the disease based on our correlation analysis and did not affect the ability of specific biomarkers to identify samples collected from real world settings. A relatively large congenital deficiency of this study is the retrospective study. Many variables that have an impact on the results are not controlled, such as comorbid anxiety, severity of psychiatric symptoms, use of psychotropic medications, etc., will directly affect those blood indicators. (3) Data of only one site was used for experiments in this study, although the sample distributions were representative as discussed above, the model needs to be validated on more samples from different settings in the next step. (4) In addition to performances of different disease durations as analyzed here, it would worth looking into the performances and feature contributions based on other sample divisions such as socio-demographic characteristics and age of onset in the near future, to examine the model stability and feature changes from different aspects. (5) One important direction in the next step is to further study the essential mechanisms behind the ability of blood biomarkers to distinguish UPD and BPD. (6) Another important direction is to use patient longitudinal data wherever possible, to build personalized models of biomarker changes during the disease process for more accurate diagnosis and tailored interventions.

## Conclusion

There is a lack of objective features for the differential diagnosis of unipolar and bipolar depression. We investigated whether a combination of hematologic biomarkers and clinical features could accurately classify unipolar and bipolar depression using machine learning methods. Experimental results demonstrated that hematologic biomarkers and onset features are reliable information that could be easily accessible in clinical settings to improve diagnostic accuracy.

## Data Availability

All data produced in the present study are available upon reasonable request to the authors

